# Characteristics and outcome of SARS-CoV-2 infection in cancer patients

**DOI:** 10.1101/2020.05.14.20101576

**Authors:** Clémence Basse, Sarah Diakite, Vincent Servois, Maxime Frelaut, Aurélien Noret, Audrey Bellesoeur, Pauline Moreau, Marie-Ange Massiani, Anne-Sophie Bouyer, Perrine Vuagnat, Sandra Malak, François-Clément Bidard, Dominique Vanjak, Irène Kriegel, Alexis Burnod, Geoffroy Bilger, Toulsie Ramtohul, Gilles Dhonneur, Carole Bouleuc, Nathalie Cassoux, Institut Curie COVID Group, Xavier Paoletti, Laurence Bozec, Paul Cottu

## Abstract

**Background:** Concerns have emerged about the higher risk of fatal COVID-19 in cancer patients. In this paper, we review the experience of a comprehensive cancer center.

**Methods:** A prospective registry was set up at Institut Curie at the beginning of the COVID-19 pandemic. All cancer patients with suspected or proven COVID-19 were entered and actively followed for 28 days.

**Results:** Among 9,842 patients treated at Institut Curie between mid-March and early May 2020, 141 (1.4%) were diagnosed with COVID-19, based on RT-PCR testing and/or CT-scan. In line with our case-mix, breast cancer (40%) was the most common tumor type, followed by hematological and lung malignancies (both 13%). Patients with active cancer therapy or/and advanced cancer accounted for 88% and 69% of patients, respectively. At diagnosis, 79% of patients had COVID-19 related symptoms, with an extent of lung parenchyma involvement ≤50% in 90% of patients. Blood count variations and C-reactive protein elevation were the most common laboratory abnormalities. Antibiotics and antiviral agents were administered in 48% and 7% of patients, respectively. At the time of analysis, 26 patients (18%) have died from COVID-19, and 81 (57%) were cured. Independent prognostic factors at the time of COVID-19 diagnosis associated with death or intensive care unit admission were extent of COVID-19 pneumonia and decreased O_2_ saturation.

**Conclusions:** COVID-19 incidence and presentation in cancer patients appear to be very similar to those in the general population. The outcome of COVID-19 is primarily driven by the initial severity of infection rather than patient or cancer characteristics.

## Introduction

Since December 2019, the world has been faced with the emergence of Coronavirus Disease 2019 (COVID-19), caused by a novel beta-coronavirus called severe acute respiratory syndrome coronavirus 2 (SARS-CoV-2)^1^. The first cases of pneumonia were reported in Wuhan City, China, in December 2019^2^ and were rapidly followed by worldwide dissemination. The World Health Organization (WHO) consequently declared COVID-19 a public health emergency of international concern^3^. COVID-19 has now affected more than four million people all over the world and has already been responsible for more than 250,000 deaths^4^. France has the 6^th^ highest number of cases of COVID-19 in the world, with more than 170,000 confirmed cases and almost 27,000 deaths^4^. The Paris area (Ile-de-France) is the epicenter of the epidemic in France^5^.

Cancer patients could represent a high-risk population in this global crisis. Based on a series of 44,672 confirmed cases in China, Wu *et al*. reported a 2.3% death rate in the general population versus 5.6% among cancer patients^6^. Several studies have simultaneously reported that pre-existing comorbidities (including cancer) constitute a risk factor associated with severe COVID-19 infection and ICU admission^7,8^. COVID-19 appears to be more frequent among cancer patients, with an over-representation of cancer patients in the confirmed COVID-19 population compared to the general population in China^9,10^. Initial retrospective cohorts of cancer patients with COVID-19 in China were characterized by a high rate of severe presentation (39% to 54% of cases) and a high mortality rate (13.5% to 29%)^10,11^. However, more recent Chinese reports on hospitalized cancer patients have shown that the average mortality rate of COVID-19 among cancer patients was closer to 5–10%^12,13^, and that patients with lung cancer or lung metastases were more prone to die from COVID-19^12^. A report from New York City suggested that only cancer patients younger than 50 were at higher risk of death when in intensive care unit^14^. The United Kingdom National Health Service reported a very weak association between cancer and the risk of dying from COVID-19^15^. Altogether, these cohorts appeared to be heavily biased as they only included hospitalized patients and were therefore unlikely to reflect the real clinical outcome of COVID-19 in cancer patients.

The first proven case of COVID-19 in France was reported on January 24, 2020. The French hospital emergency response plan (*“Plan Blanc”*) and a nation-wide lockdown were implemented on March 6, 2020. Institut Curie is a prominent comprehensive cancer center with three sites in Paris, Saint Cloud and Orsay (Supplementary Figure S1). In view of the urgent need for more data on COVID-19 in cancer patients, we set up an institutional registry at the very beginning of the pandemic in France in order to: (i) more accurately determine the incidence of the disease, (ii) more precisely understand the outcomes in this fragile population, and (iii) identify specific risk factors for poor outcome. We prospectively registered all patients with confirmed or highly suspected COVID-19, including ambulatory patients with specific follow-up. From March 13 to May 1, 2020, 9,842 cancer patients were referred at one of the Institut Curie facilities (including 7,833 patients receiving active therapy) for any type of cancer, including breast cancer (45%), eye cancer (9%), head and neck cancer (6%), urological cancer (6%), gynecological cancer (6%), gastrointestinal cancer (6%), hematological malignancy (6%), sarcoma (5%), and lung cancer (5%). In this paper, we report the clinical characteristics and outcomes of SARS-CoV-2-infected patients treated or followed for cancer at Institut Curie.

## Patients and Methods

### Registry implementation and data collection

An institutional database was implemented to prospectively collect information concerning cancer patients treated or followed at Institut Curie with suspected or confirmed SARS-CoV-2 virus infection. This registry was approved by the Institut Curie Institutional Review Board (IRB). No documentation or informed consent was required according to French regulations, as this study was strictly observational. Redcap® software was used as data repository. Starting on March 13, we consecutively registered patients with symptoms suggestive of COVID-19, and/or confirmed COVID-19 by nasopharyngeal swab RT-PCR, and/or images suggestive of COVID-19 on chest CT scan. The design and reporting of this case-cohort study strictly complied with STROBE guidelines^16^.

The main data recorded were as follows: cancer history (age, sex, primary tumor site, localized or advanced tumor); current cancer treatment intent according to 3 categories (curative intent for adjuvant/neoadjuvant/early-stage treatment, disease control when active cancer treatment was administered for advanced cancer, and palliative phase); COVID-19 features (symptoms, nasopharyngeal swab RT-PCR result, when performed, suggestive chest CT scan images, when performed, laboratory parameters); COVID-19 severity (ambulatory patients, hospitalization, transfer to intensive care unit (ICU), COVID-19- or cancer-related death); consequences on current cancer treatment (delayed, canceled, modified). Comorbidities and comedications were also collected. Patients who received noninvasive respiratory support in conventional hospitalization wards were considered to be hospitalized patients and not as ICU patients. Treatments specifically administered for COVID-19 were also recorded. Systematic medical follow-up was performed at day 7, day 14 and day 28 after inclusion in the registry. Ethnic group analyses are not allowed per French regulations.

### COVID-19 diagnosis

From March 13, 2020, a systematic screening comprising a questionnaire and systematic body temperature check was implemented at entrance of all our facilities. According to national guidelines, SARS-CoV-2 RNA testing on nasopharyngeal swab (RT-PCR testing) was initially restricted to healthcare workers and severely ill patients^17^. Tests became more readily available (upon prescription) after March 25, and patients with COVID-19 symptoms were then tested for SARS-CoV-2 RNA whenever possible. SARS-CoV-2 RNA testing also became mandatory for all patients, even asymptomatic, scheduled for surgery at Institut Curie. Cancer care, imaging and laboratory assessments are detailed in Supplementary Methods.

### Eligibility

Registry inclusion criteria were: patients either systematically screened for SARS-CoV-2 infection, or currently under treatment or surveillance at the Institut Curie for a solid tumor or hematological malignancy, presenting any symptom of COVID-19, or with confirmed COVID-19 by nasopharyngeal swab RT-PCR, or presenting chest CT scan images suggestive of COVID-19. Only patients with positive imaging and/or positive RT-PCR testing were considered positive for COVID-19 for the present analysis, regardless of their symptoms.

### Study objectives

The primary objectives of the study were to evaluate the incidence of COVID-19 in Institut Curie patients and describe the clinical presentation, management and outcome of COVID-19 during the 28 days following onset of the first symptoms. Secondary objectives were to evaluate the impact of COVID-19 on cancer care, and factors associated with COVID-19 severity, such as concomitant treatments or comorbidities.

### Statistics

Descriptive statistics are reported in all COVID patients and in the subgroup with both positive RT-PCR and chest CT-scan. The main clinical outcome was defined as death due to COVID-19 and/or ICU admission during follow-up. Univariate and multivariate prognostic factor analysis was performed. For the multivariate analysis, a backward selection model was applied starting from variables significant at the 10% level with less than 10% of missing data. Sensitivity analyses were performed on the prognostic factors for death alone and for time to worsening (ICU or death) using log-rank test and semi-parametric model after checking the proportional hazard assumption. All analyses were performed with SAS v9.4 software.

## Results

### Population and symptoms

From March 13, 2020, to April 25, 2020, 186 patients were entered into the registry. Forty-five patients were subsequently excluded due to uncertain diagnosis, leaving 141 patients with positive RT-PCR and/or positive chest CT-scan for the final analysis (Figure 1). The weekly number of tested patients is illustrated in Supplementary Figure S2. Considering the 9,842 patients who attended the Institut Curie at least once during the same period, the minimum estimated incidence of COVID-19 in our patients was 1.4%. Patient characteristics are detailed in Table 1, pointing some numerical differences according to initial presentation. Overall, median age was 62 years, and 37 (26%) patients were older than 70. Of note, 25 (18%) patients were obese. Main pre-existing comorbidities deemed relevant in the context of COVID-19 were hypertension (34%), smoking (18%) and diabetes (17%). The most common comedications were anticoagulants (24%), angiotensin-converting enzyme inhibitors/ Angiotensin II Receptor Blockers (20%) and corticosteroids (18%).

**Figure 1.**
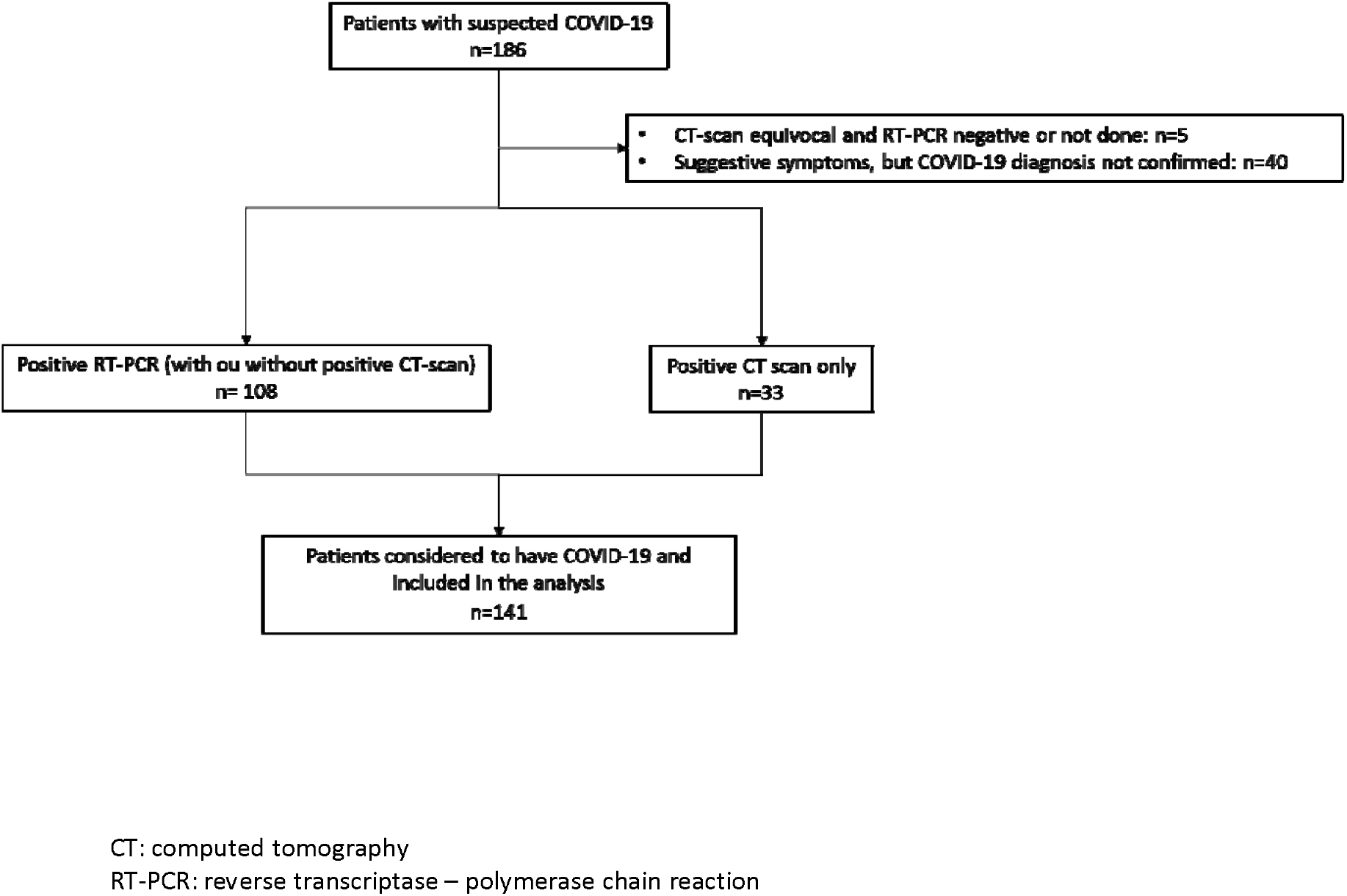
Flow chart.

**Table 1.**
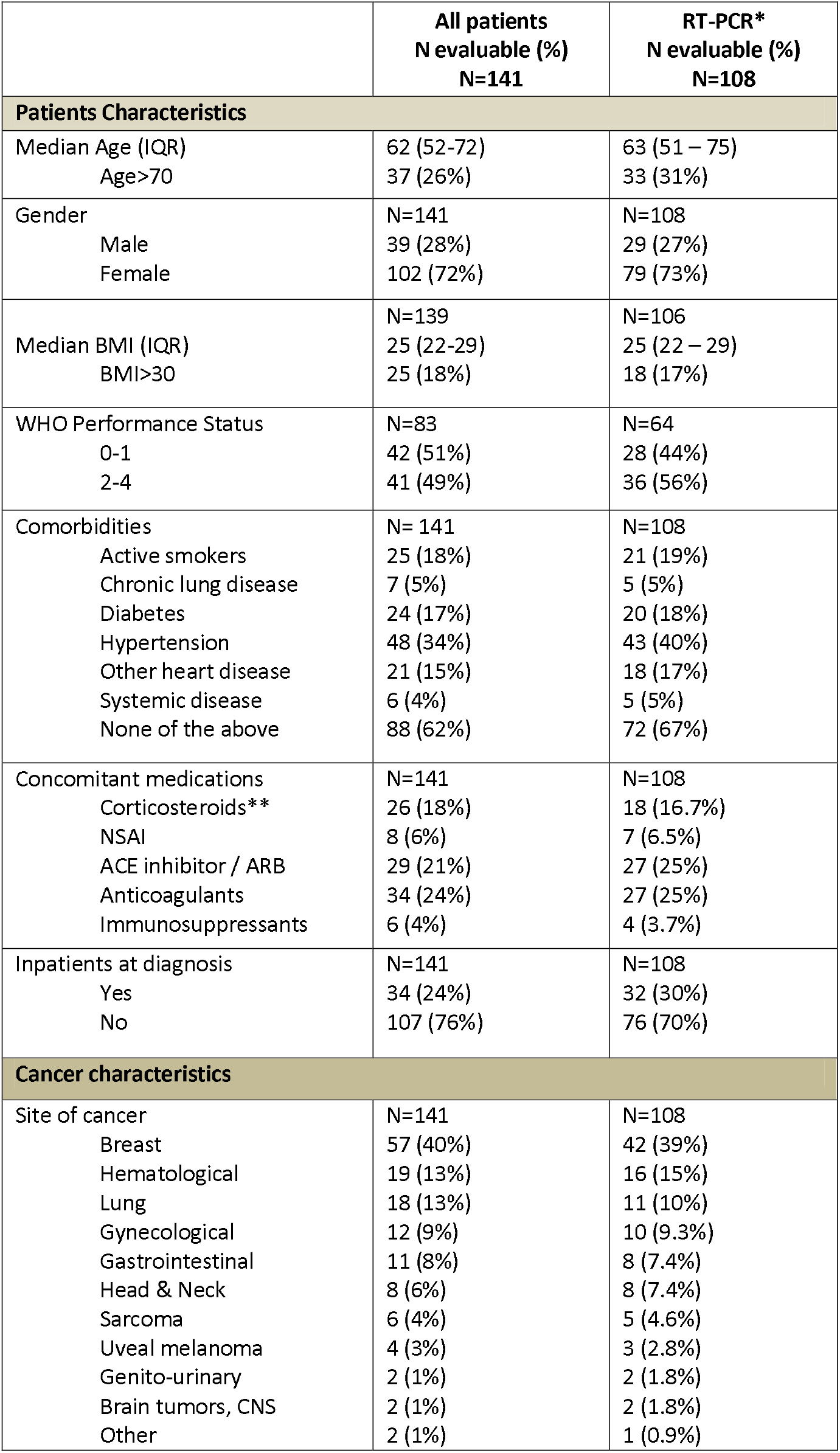
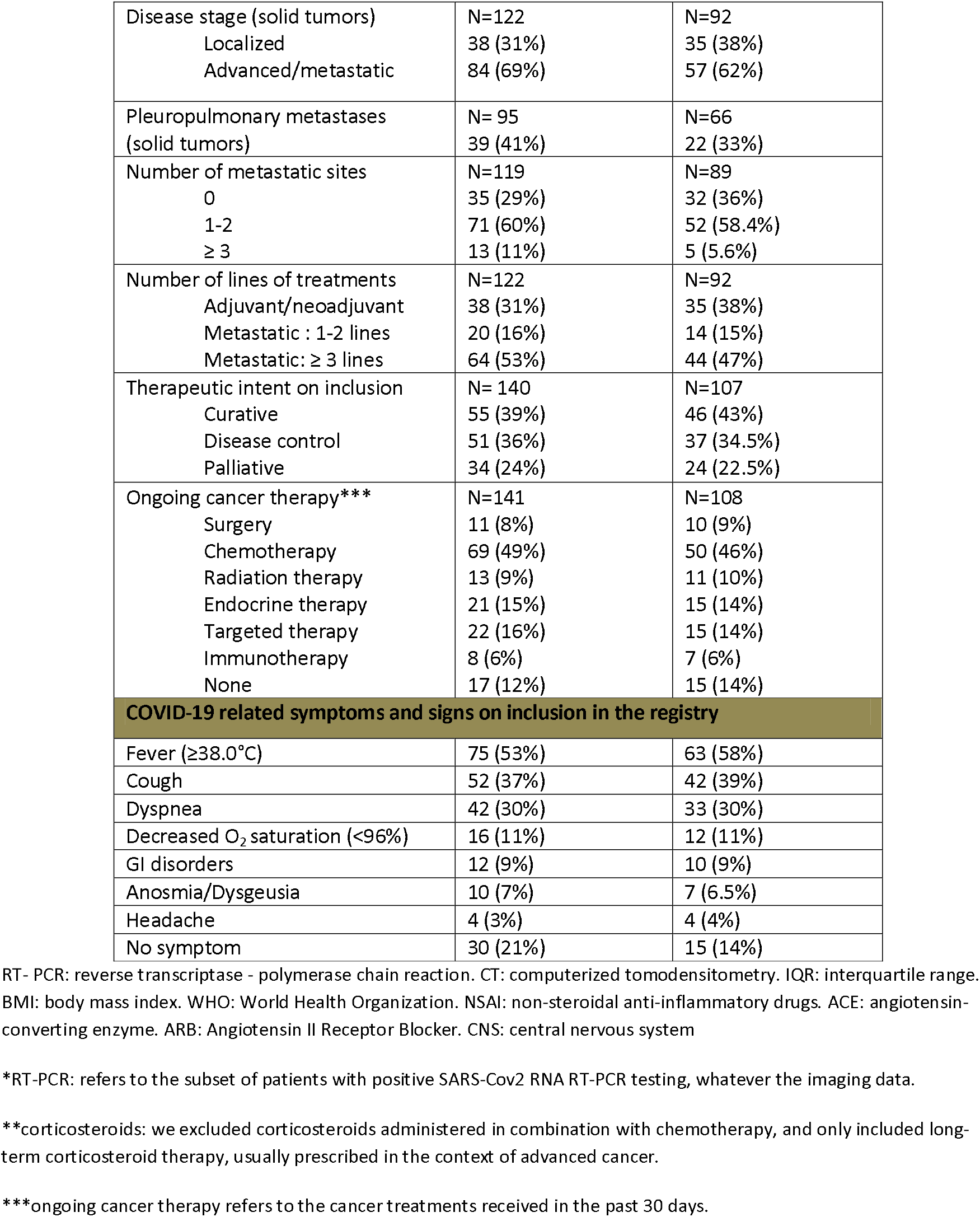
Population characteristics.

Breast cancer was the most common cancer type (40%), followed by lung cancer (13%), hematological malignancies (13%), gynecological cancers (9%), and other miscellaneous tumors in line with the Institut Curie case-mix. At the time of inclusion in the registry, 84 patients (69% of solid cancers) had advanced/metastatic cancer, including 39 patients (41%) with pleuropulmonary metastases. Seventeen patients (12%) were under surveillance for their cancer and 124 (88%) were receiving ongoing treatment. Thirty-four patients (24%) had already been hospitalized for cancer care for more than two days at the time of COVID-19 onset. Thirty-nine percent of patients received treatment with curative intent (adjuvant/neoadjuvant/early-stage), while 36% received treatment for disease control and 24% received palliative care.

COVID-19-related clinical symptoms on inclusion in the registry were observed in 111 (79%) of the 141 patients. Fever (≥38°C) and cough were the most common symptoms (53% and 37%, respectively). Dyspnea was observed in 30% of patients, including 10% with respiratory distress. Other less common symptoms are shown in table 1.

### Imaging and laboratory findings

Baseline chest CT scan findings, available for 80 patients, are described according to the extent of pulmonary parenchyma COVID-19-related alterations and clinical findings. The extent of parenchymal disease appeared to be relatively independent of the underlying patient or cancer key characteristics such as disease site or stage (Figure 2A), comorbidities and co-medications (Figure 2B), or mode of COVID-19 diagnosis (Figure 2C), although visual analysis suggested more extensive lung involvement in a small subset of patients with hematological malignancies. Detailed radiological findings are reported in Supplementary table S1. Overall, extent of lung parenchyma involvement was ≤50% in 90% of patients. Detailed radiological findings are reported in Supplementary tables S1 and S2.

**Figure 2.**
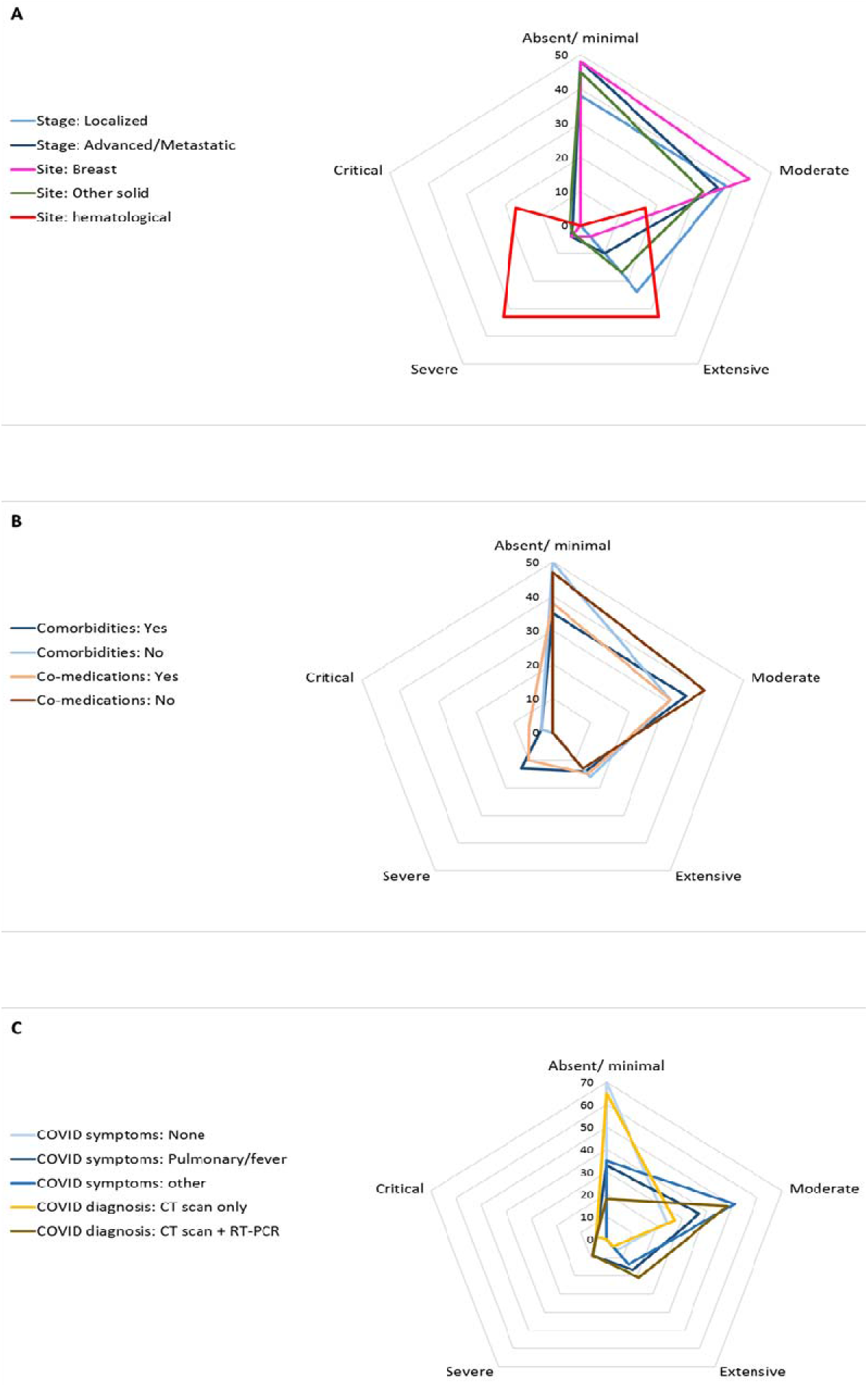
Baseline COVID-19-related findings on chest CT scan according to main baseline characteristics in 80 patients. Distribution of the initial extent of pulmonary parenchymal lesions is displayed according to cancer-related (2A), patient-related (2B) or COVID-19 diagnosis-related (2C) related parameters. Numbers indicate the percentage of each imaging pattern according to the five-class classification we retained: absent/minimal (0-<10%); moderate (10–25%); extensive (26–50%); severe (51–75%); critical (>75%). The longest spokes indicate the highest frequency of the corresponding pattern.

Laboratory test results (worse result at any time point) are detailed in Supplementary Table S2, and test results of special interest are shown in Table 2. We observed statistically significant, although clinically meaningless, differences in blood counts. Significant elevation of baseline serum C-reactive protein was recorded in 35 evaluable patients.

**Table 2.** Significant laboratory findings at COVID-19 diagnosis compared to reference laboratory values.

|  | Reference*<br>(N evaluable) median value (IQR) | At COVID-19 diagnosis<br>(N evaluable) value (IQR) | Mean of the differences<br>(N evaluable) value (IQR) | p |
| --- | --- | --- | --- | --- |
| Blood Count |  |  |  |  |
| Hemoglobin (g/dL) | (N=114) 11.6 (10.5 – 13.2) | (n=100) 11.0 (9.2-12.4) | (N=87) -1.16 (-2.3 – +0.1) | **<.0001** |
| Absolute Neutrophil count (10<sup>9</sup>/L) | (N=116) 3.93 (2.59 – 5.81) | (n=97) 4.19 (2.52-7.32) | (n=85) +0.77 (-1.72 – +1.95) | **.42** |
| Absolute Lymphocyte count (10<sup>9</sup>/L) | (N=114) 1.18 (0.79 – 1.74) | (n=96) 0.83 (0.48-1.38) | (n=82) -0.31 (-0.77 – +0.09) | **0.03** |
| Platelets (10<sup>9</sup>/L) | (N=114 ) 240 (169 – 326) | (n=100) 190 (132-280) | (n=87) -38.6 (-100 – +26) | **0.018** |
| CRP (mg/L) | (N=46) 20 (4.6-53.2) | (N=71) 84 (41-142) | (N=35) +72 (+11 – +118) | **0.0001** |

CRP: C-reactive protein.
\*reference =laboratory test results obtained no more than 2 months before inclusion in the registry.

### COVID-19 therapy and clinical outcome

Patients were either discharged home (64%), or hospitalized (36%) depending on the severity of the symptoms. Antibiotics were prescribed in about 50% of patients according to local guidelines. Antiviral drugs were administered in selected cases after a thorough review of the patient’s medical history (Supplementary Table S3). Most patients stayed at home and remained asymptomatic or very slightly symptomatic between baseline and Day 28 (Figure 3). However, 11 (8%) patients were transferred to the intensive care unit at some time during the course of the disease and, at the time of analysis, 26 patients (18%) had died from COVID-19, 4 had died from terminal cancer, 31 (22%) were recovering and 81 (57%) were cured. Lung cancers had the worst outcome (n = 6 i.e. 23% of all deaths) followed by hematological malignancies (n = 5, 19%), breast (n = 5, 19%) and gynecological cancers (n = 2, 7%). Twenty patients (77% of deceased patients) had received more than 3 lines of treatment for metastatic disease. The place of death was home (1), Institut Curie or other hospitals (12 and 8 respectively) and was missing for five patients.

**Figure 3.**
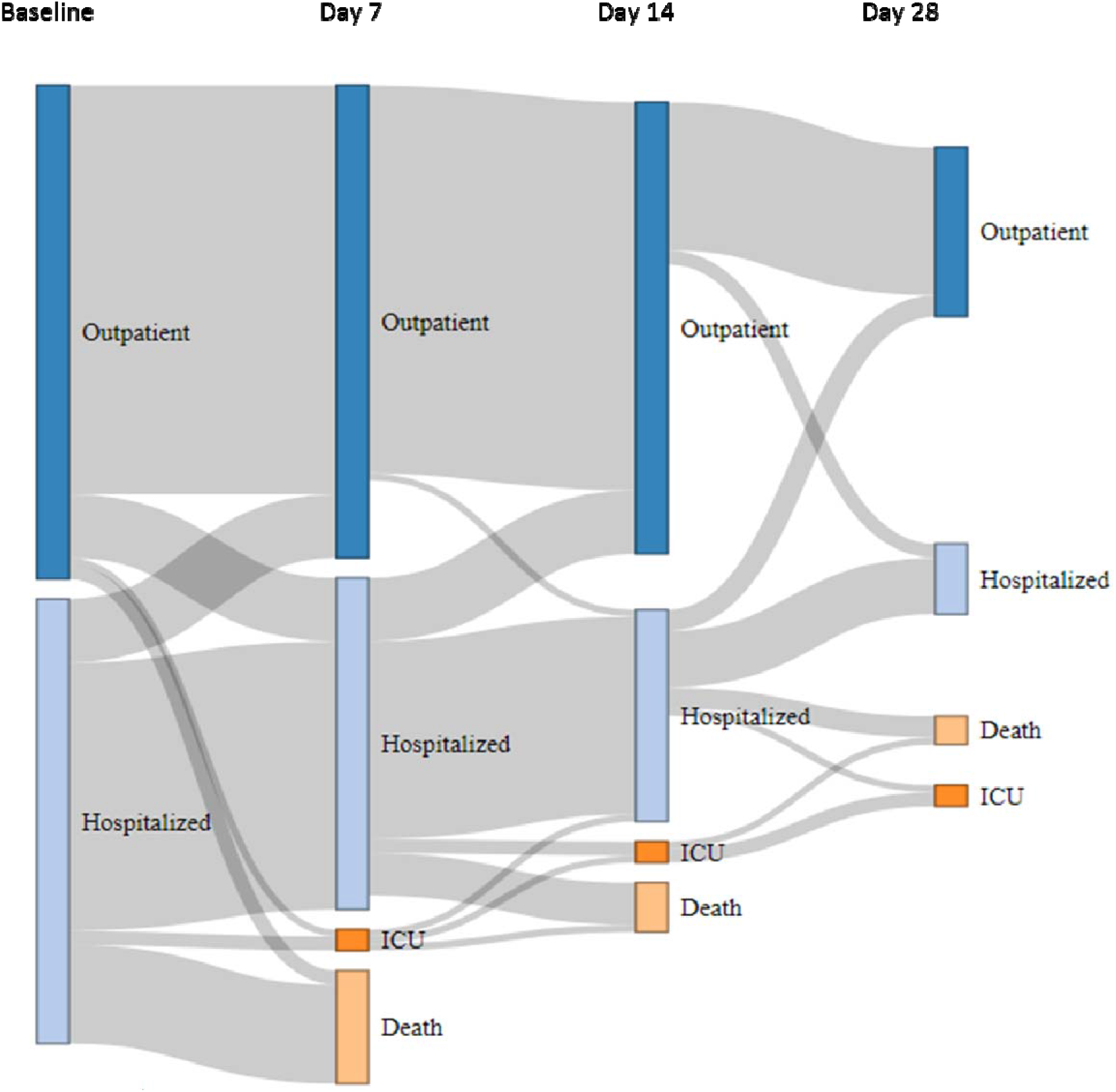
Patient trajectory from baseline to day 28. Diagram showing the changing status of cancer patients with COVID-19, based on individual follow-up. At baseline, patients were either immediately discharged home (dark blue bars) or hospitalized (light blue bars), including patients already hospitalized at the time of COVID-19 diagnosis (hospital-acquired infection). The patient’s status may have changed on Days 7, 14 and 28 to one of the following four modalities: discharged home (dark blue bars), hospitalized (light blue bars), admitted to ICU (dark orange bars), and deceased (light orange bars). The grey flows between bars are proportional to the number of patients at each step

A total of 35 patients had poor outcome (transfer to ICU or death). On univariate analysis (Table 3), factors related to patient (Age > 70, male gender), cancer (non-breast cancer, palliative setting), or SARS-CoV-2 infection (pulmonary symptoms, extent of radiological signs of pneumonia) were significantly associated with an increased risk of poor outcome. A particularly interesting finding on multivariate analysis was that only a baseline decrease in O_2_ saturation and the extent of radiological signs of lung damage remained predictors of death and/or ICU admission, with odds ratios (OR) of 6.7 (95% CI: 1.5 to 3.0) and 2.5 (95% CI: 1.3 to 4.8), respectively. The ORs for age (>70) and non-breast cancer were similar on univariate and multivariate analyses, but were no longer statistically significant. Sensitivity analyses of the risk of death alone and for the time to death or ICU admission provided the same significant associations.

**Table 3.** Analysis of prognostic factors associated with either ICU admission or death.

| Adverse prognostic factors | Univariate |  |  | Multivariate |  |  |
| --- | --- | --- | --- | --- | --- | --- |
|  | OR | 95%CI | p value | OR | 95%CI | p value |
| **Patient-related parameters** |  |  |  |  |  |  |
| Age >70 (vs ≤ 70) | **2.4** | 1.1 - 5.5 | **0.04** | 2.4 | 0.6 – 9.7 | 0.21 |
| WHO performance status 2+ (vs 0-1) | **4** | 1.5 – 10.8 | **<0.01** |  |  |  |
| BMI ≥30 (vs <30) | 0.5 | 0.2 – 1.6 | 0.25 |  |  |  |
| Male (vs Female) | **2.6** | 1.1 – 5.8 | **0.02** |  |  |  |
| Any comorbidity Yes (vs No) | 1.2 | 0.5 – 2.7 | 0.64 |  |  |  |
| Hypertension Yes (vs No) | 1.9 | 0.9 – 4.3 | 0.10 |  |  |  |
| Smoking Yes (vs No) | 0.9 | 0.3 – 2.6 | 0.92 |  |  |  |
| Concomitant medications Yes (vs No) |  |  |  |  |  |  |
| Corticosteroids | 1.45 | 0.6 – 3.7 | 0.43 |  |  |  |
| NSAID | 0.42 | 0.1 – 3.5 | 0.42 |  |  |  |
| ACE inhibitor / ARB | 1.49 | 0.6 – 3.7 | 0.48 |  |  |  |
| Anticoagulants | 1.99 | 0.9 – 4.6 | 0.11 |  |  |  |
| Immuno-suppressants | 3.2 | 0.6 – 17 | 0.16 |  |  |  |
| In patients infection (vs outpatient) | 1.1 | 0.5 – 2.7 | 0.80 |  |  |  |
| **Cancer-related parameters** |  |  |  |  |  |  |
| Site |  |  |  |  |  |  |
| Breast (ref.) | 1 |  |  |  |  |  |
| Non breast | **3.6** | 1.4 – 8.9 | **<0.01** | 3.6 | 0.8 – 17.1 | 0.1 |
| Lung | **3.6** | 1.1 – 12.6 |  |  |  |  |
| Other solid tumor | **4.5** | 1.3 – 15.6 |  |  |  |  |
| Hematological malignancy | **3.3** | 1.2 – 8.2 |  |  |  |  |
| Cancer stage (solid tumors) |  |  |  |  |  |  |
| Advanced /metastatic (vs localized) | **3.2** | 1.0 – 10.3 | **0.05** |  |  |  |
| Pleuropulmonary metastasis (Solid tumors) |  |  |  |  |  |  |
| Yes (vs No) | 1.0 | 0.4 – 2.8 | 0.98 |  |  |  |
| Therapeutic intent |  |  |  |  |  |  |
| Curative (ref.) | 1 |  |  |  |  |  |
| Control | 1.8 | 0.7 – 4.9 | **0.01** |  |  |  |
| palliative | **4.6** | 1.7 – 12.7 | (global) |  |  |  |
| Line of treatment |  |  |  |  |  |  |
| Neoadjuvant/Adjuvant (ref.) | 1 |  |  |  |  |  |
| Metastatic ≤ 2 lines | 2.1 | 0.5 – 9.5 | 0.02 |  |  |  |
| Metastatic 3+ lines | 4.5 | 1.4 – 14.0 | (global) |  |  |  |
| **COVID-19-related parameters** |  |  |  |  |  |  |
| Symptoms |  |  |  |  |  |  |
| Dyspnea Yes (vs No) | **4.3** | 1.9 – 9.6 | **<0.01** |  |  |  |
| Fever Yes (vs No) | 1.7 | 0.8 – 3.7 | 0.19 |  |  |  |
| SpO<sub>2</sub> <96% Yes (vs No) | **3.6** | 1.3 – 10.6 | **0.02** | **6.7** | 1.4 – 30 | **0.01** |
| Radiology |  |  |  |  |  |  |
| % alteration (per category) | **2.4** | 1.3 – 4.2 | **0.003** | **2.5** | 1.3 – 4.8 | **0.004** |

OR: odds ratio. CI: confidence interval. WHO: World Health Organization. BMI: body mass index. NSAID: non-steroidal anti-inflammatory drugs. SpO<sub>2</sub>: oxygen saturation as detected by the pulse oximeter

### Impact on cancer care

Based on individual assessment, chemotherapy, targeted therapy or immune checkpoint inhibitor therapy were delayed or stopped in 68%, 67% and 83% of cases, respectively. Planned surgical procedures were postponed in 79% of cases. Ongoing radiation therapy was delayed or canceled in 69% of cases. Elective surgery was rescheduled after a mean of 3 weeks (15 days to 6 weeks).

## Discussion

The COVID-19 pandemic has and continues to have a dramatic impact on health care systems worldwide. Cancer patients potentially represent one of the most vulnerable patient populations, and continuously updated data on how COVID-19 affects cancer care are essential. To the best of our knowledge, we report the first comprehensive prospective cohort of cancer patients, not confined to hospitalized patients, but also including outpatients with systematic and dedicated follow-up. We report three types of results.

Firstly, with 141 patients with COVID-19 observed during the period from mid-March to early May, the minimum incidence rate of SARS-CoV-2 infection in the overall cohort of patients treated at Institut Curie during this period was 1.4%, (or 1.7% when only considering patients with active cancer treatment). Although this incidence rate is potentially underestimated, it is not higher than the 5.7% rate estimated for the whole French population during the same period with the same diagnostic guidelines, i.e. clinical screening, RT-PCR on naso-pharyngeal swabs and chest CT scan^18^. Furthermore, the proportion of infected people in the Paris area (“Ile-de-France”) in which the Institut Curie facilities are located (Supplementary Figure S1) is estimated to be 12.3%^18^, clearly suggesting that cancer patients, as a whole, are not more prone to SARS-CoV-2 infection than the general population when strict lockdown measures are implemented.

Secondly, the clinical features of COVID-19 in cancer patients appear to be very similar (Table 1, Figure 2) to those observed in the general population^6,7,11^. The vast majority of patients were symptom-free and had been discharged home by Day 28 (Figure 3), and no specific radiological (Figure 2, Table S3) or laboratory (Tables 2 and S2) findings were observed. Furthermore, the mortality rate observed in our prospective cohort (19%) was also similar to that reported in the general population (up to 20%)^19^, suggesting that the population of cancer patients presents a similar risk of dying from COVID-19 to that of the general population, in contrast with the results of several early retrospective studies^8,10,11,14^. This in line with the NHS report showing a slight increase of dying from COVID-19 limited to inpatients with recent diagnosis of cancer^15^. One of the most striking observations in our cohort is that, when using ICU admission and/or death as a relevant clinical endpoint, COVID-19-related characteristics clearly appeared to be the most powerful predictors of poor outcome, as partly reported in several retrospective general cohorts^2,8,11^. We observed a potential trend for a higher risk of death/ICU admission in older patients, and in patients with non-breast cancer (mostly lung and hematological malignancies) or in the palliative setting. Inpatients diagnosed with COVID-19 had not a higher risk of death from the infection. Several hypotheses could also explain the lower than expected rate of severe COVID-19. In this study, in addition to hospitalized patients, we also included ambulatory patients, corresponding to patients with milder symptoms and a more favorable disease outcome. Of note, 24% of the patients of this cohort received anticoagulants, which may have prevented thromboembolic complications^20^. We did not observe any case of cytokine storm syndrome^21^, which may have been prevented by pre-existing inflammatory state^22^, or conversely immune exhaustion^23^.

Thirdly, we also prospectively recorded the impact of COVID-19 on cancer care. More than two-thirds of scheduled cancer treatments were canceled, postponed or significantly modified. In a separate study, we evaluated that first consultations for breast cancer decreased by 34% compared to the same period in 2019^24^. As highlighted by many authors, the COVID-19 pandemic will very likely have a major collateral impact on cancer outcome in the next months to come^25–29^, while placing the cancer workforce under a considerable strain^30^.

There are limitations to this study. Access to RT-PCR tests differed between the beginning and the end of the cohort registration period, due to limited test availability at the beginning of the registration period (Supplemental Figure S2). It is far too early to evaluate the actual cancer outcome of cancer patients in the COVID-19 era. We are also aware that breast cancer patients may be overrepresented in our cohort, which could contribute to the overall good prognosis of SARS-CoV-2 infection observed in this study. However, our practice is similar to that of other comprehensive cancer centers, and our experience can therefore provide useful information in this specific context. This registry needs to be continued in order to provide more extensive and more robust data.

In summary, early reports on cancer and COVID-19, mostly based on hospitalized cancer patients, tended to suggest that cancer patients were more prone to suffer from SARS-CoV-2 infection and experienced a higher mortality rate. Special attention must be paid to patients with lung cancer and hematological malignancies. However, risk factors for COVID-19 mortality are mostly infection-related rather than cancer-related. Overall, our findings from a large prospective cohort of representative cancer patients and treatments, including ambulatory, non-hospitalized patients, strongly suggest that COVID-19 is neither more frequent nor more fatal in cancer patients as a whole. A simple baseline clinical assessment combining O_2_ saturation measure and a chest CT scan clearly identifies patients at risk of poor outcome and should be widely recommended in cancer patients with suspected COVID-19. As of April 28, a systematic serological testing is now performed in all patients (those included in the present cohort as well as new patients). We believe that the data presented here, combined with more accurate epidemiological data based on longer follow-up and generalized antibody testing, will pave the way to safer and individualized care and lockdown ease for cancer patients in the COVID-19 era.

## Data Availability

Availability of data and materials
The data underlying this article cannot be shared publicly due to current French HIPAA regulations (birthdate, admission date, discharge date, date of death). Data will be shared on reasonable request to the corresponding author.

## Ethics approval

The COVID-19 registry was approved by the Institut Curie internal review board,which waived the need for informed consent, due to the observational nature of this registry.

## Availability of data and materials

The data underlying this article cannot be shared publicly due to current French HIPAA regulations (birthdate, admission date, discharge date, date of death). Data will be shared on reasonable request to the corresponding author.

## Authors’ contributions

CB, SD, VS, MF, AN, AB, PM, MAM, ASB, PV, SM, FCB, DV, TR, XP, LB and PC contributed to data collection and interpretation. XP, LB and PC set up the registry and contributed to data interpretation. CB, SD, MF, AN, and FCB contributed to manuscript writing. CB, SD, XP, LB and PC collected data, contributed to the analysis and wrote the manuscript. XP performed the statistical analyses. All authors read and approved the final manuscript.

## Acknowledgments

Institut Curie COVID Group: Elisabeth Angellier, Muriel Belotti, Sara Boubakeur, Hervé Brisse, Bruno Buecher, Sylvie Carrié, Laetitia Chanas, Pascal Chérel, Gilles Créhange, Christelle Colas, Olivier Collin, Baudouin Courtier, Hélène Delhomelle, Emmanuelle Fourme, Thomas Frédéric-Moreau, Pierre Fumoleau, Marion Gauthier-Villars, Jean-David Heisbourg, Sophie Lassalle, Marine Le Mentec, Jane Muret, Sophie Métivier, Antoine de Pauw, Jean-Yves Pierga, Mael Priour, Roman Rouzier, Mary Saad, Claire Saule, Dominique Stoppa-Lyonnet, Anne Tardivon, Anne Vincent-Salomon.

